# Estimating maximal oxygen consumption from heart rate response to submaximal ramped treadmill test

**DOI:** 10.1101/2020.02.18.20024489

**Authors:** Tomas I. Gonzales, Kate Westgate, Stefanie Hollidge, Tim Lindsay, Justin Jeon, Søren Brage

**Author notes:** Address for correspondence: Dr. Søren Brage, MRC Epidemiology Unit, University of Cambridge, School of Clinical Medicine, Box 285, Institute of Metabolic Science, Cambridge Biomedical Campus, Cambridge CB2 0QQ, United Kingdom.

## Abstract

The Cambridge Ramped Treadmill Test (CRTT) is an incremental, multistage exercise test (I: steady-state walk, II: walk ramped speed, III: walk ramped incline, and IV: run ramped speed on flat). It is typically deployed as a submaximal test with flexible test termination criteria, making it an attractive option for population-based studies of cardiorespiratory fitness. We conducted a study in healthy adults to test the validity of maximal oxygen consumption estimates (VO_2_ max; ml O_2_·kg^-1^·min^-1^) predicted from CRTT heart rate response using several methods: a heart rate-to-work rate linear regression method across several test termination criteria, either when a percentage of age-predicted maximal heart rate was achieved (50% through 100%) or at the end of distinct CRTT stages (II, III, and IV); and two single-point walk-test calibration methods using data from either CRTT stage I (low-point method) or stage II (high-point method). For estimates from the linear regression method, prediction bias ranged from −3.0 to −1.6 ml O_2_·kg^-1^·min^-1^ and Pearson’s *r* ranged from 0.57 to 0.79 for endpoints at percentages of age-predicted maximal heart rate; results were similar for stages III and IV endpoints, but predictions using data only up to stage II had poorer agreement. Agreement was moderate when using the low-point (mean bias: −4.3 ml O_2_·kg^-1^·min^-1^; Pearson’s *r*: 0.71) and high-point (mean bias: −3.5 ml O_2_·kg^-1^·min^-1^; Pearson’s *r*: 0.69) methods. Heart rate response to the CRTT can be used to predict VO_2_max with acceptable validity in common epidemiological scenarios.

## Introduction

For continuous incremental exercise at submaximal intensities, statistical techniques like linear regression can be used to characterise work rate (WR) as a function of heart rate (HR) _1_. HR increases in parallel with oxygen consumption (VO_2_) and reaches a maximal value (HRmax) when maximal oxygen consumption (VO_2_max) is achieved ^2^. Assuming a linear relationship between HR and WR, which holds at maximal intensities ^3,4^, VO_2_max can be predicted by: 1) extrapolating the HR-to-WR regression line to age-predicted HRmax ^5^; 2) converting the extrapolated WR value to net VO_2_ using a caloric equivalent for oxygen ^6^; and 3) adding an estimate of resting energy expenditure (REE). The validity of predicted VO_2_max values would depend on sources of error that arise at each step of this approach. If the range of submaximal WR values is too narrow or elicits an inadequate HR response, the HR-to-WR regression line may diverge from measured VO_2_ dynamics when extrapolated to maximal intensities, biasing VO_2_max predictions. Random error associated with estimating HRmax from age will shift the extrapolation endpoint from the participant’s true HRmax, decreasing VO_2_max prediction precision ^7^. Finally, the estimation of REE may be biased by using a common REE value across all individuals (i.e. 1 metabolic equivalent; MET)^8^ as covariates such as age, sex, and body composition are not adjusted for ^9,10^.

In this study, we examine the validity of several methods for predicting VO_2_max from HR response to the Cambridge Ramped Treadmill Test (CRTT). The CRTT is an incremental, multistage, submaximal exercise test, originally designed for individual calibration of HR-to-energy expenditure for the quantification of physical activity energy expenditure during free-living ^11,12^. The CRTT also has utility for estimation cardiorespiratory fitness in population-based studies, the interpretation of which would depend on its validity and has yet to be examined. Study participants completed an extended version of the CRTT to exhaustion while WR, HR, and VO_2_ were recorded. Using this design, we report the validity of inferences from different testing conditions by comparing VO_2_max values predicted from data available at several test endpoints with directly measured VO_2_max. We then provide recommendations for the interpretation of CRTT data for population-based studies of cardiorespiratory fitness.

## Methods

### Participants

Participants were recruited via poster adverts in a local hospital in Cambridge, UK, and written invitation to participants from the Fenland study, an ongoing population-based cohort study ^13^. Exclusion criteria included prevalent diabetes, pregnancy or lactation, inability to walk unaided, psychosis, and terminal illness. Participants underwent a medical screening prior to testing, consisting of a health assessment questionnaire, review of medications, resting electrocardiogram, and blood pressure measurement. All volunteers provided written informed consent, and the study was approved by the Cambridgeshire Research Ethics Committee (Ref:07/Q0106/21).

### Study procedure

Participants abstained from eating, drinking (except water), smoking, and exercise for at least 2 hours prior to testing. Height was measured with a rigid stadiometer (SECA 240; Seca, Birmingham, UK). Weight and fat-free mass were measured in light clothing with calibrated scales (TANITA model BC-418 MA; Tanita, Tokyo, Japan). Resting HR (RHR) was measured with the participant in a supine position using a combined HR and movement sensor (Actiwave Cardio, CamNtech, Cambridge, UK) attached to the chest at the base of the sternum by two standard ECG electrodes. HR was recorded for 15 minutes and RHR was calculated as the mean HR measured during the last 3 minutes. Participants wore another combined sensor (Actiheart, CamNtech, Cambridge, UK) for 1 week of free-living to measure sleeping HR (SHR) ^14^. HR above SHR (HRaS) was calculated as HR minus SHR.

### VO_2_ and energy expenditure

VO_2_ during rest and during the CRTT was measured using a computerised metabolic cart (Oxycon Pro, Erich Jaeger GmbH, Hoechberg, Germany); this system has been validated previously ^15^. Energy expenditure was calculated by indirect calorimetry according to Weir ^16^. REE was measured in the supine position with a ventilated hood positioned over the participant’s head for 15 minutes; the mean of the last 5 min was used in analysis.

### Cambridge Ramped Treadmill Test (CRTT)

As described in detail elsewhere ^12^, the CRTT has four sequential stages: Stage I) steady-state walk; walking at 3.2 km·h^-1^ at 0% incline for 3 min, Stage II) walk ramped speed; walking while treadmill speed increases from 3.2 to 5.2 km·h^-1^ at 0% incline for 6 min, Stage IIIa) walk ramped incline, first subphase; walking at 5.2 km·h^-1^ while incline increases from 0 to 6% for 3 min, Stage IIIb) walk ramped incline, second subphase; walking while treadmill speed increases from 5.2 to 5.8 km·h^-1^ and incline from 6 to 10.2% for 3 min, Stage IVa) run ramped speed on flat, first subphase; treadmill speed increases from 5.8 to 9.0 km·h^-1^ and incline decreases to 0% for 1 min, and Stage IVb) run ramped speed on flat, second subphase; running while treadmill speed increases from 9.0 to 12.6 km·h^-1^ and at 0% incline for 4.5 min. The test ended with a 2-minute standing recovery period. Instantaneous WR values, expressed as physical activity intensity (PAI, J·kg^-1^·min^-1^), were computed from treadmill speed and incline according to measured PAI in Brage et al. 2007 ^12^. Figure 1 provides a graphic representation of the CRTT and exemplar data demonstrating change in WR and HR across CRTT stages.

**Figure 1.**
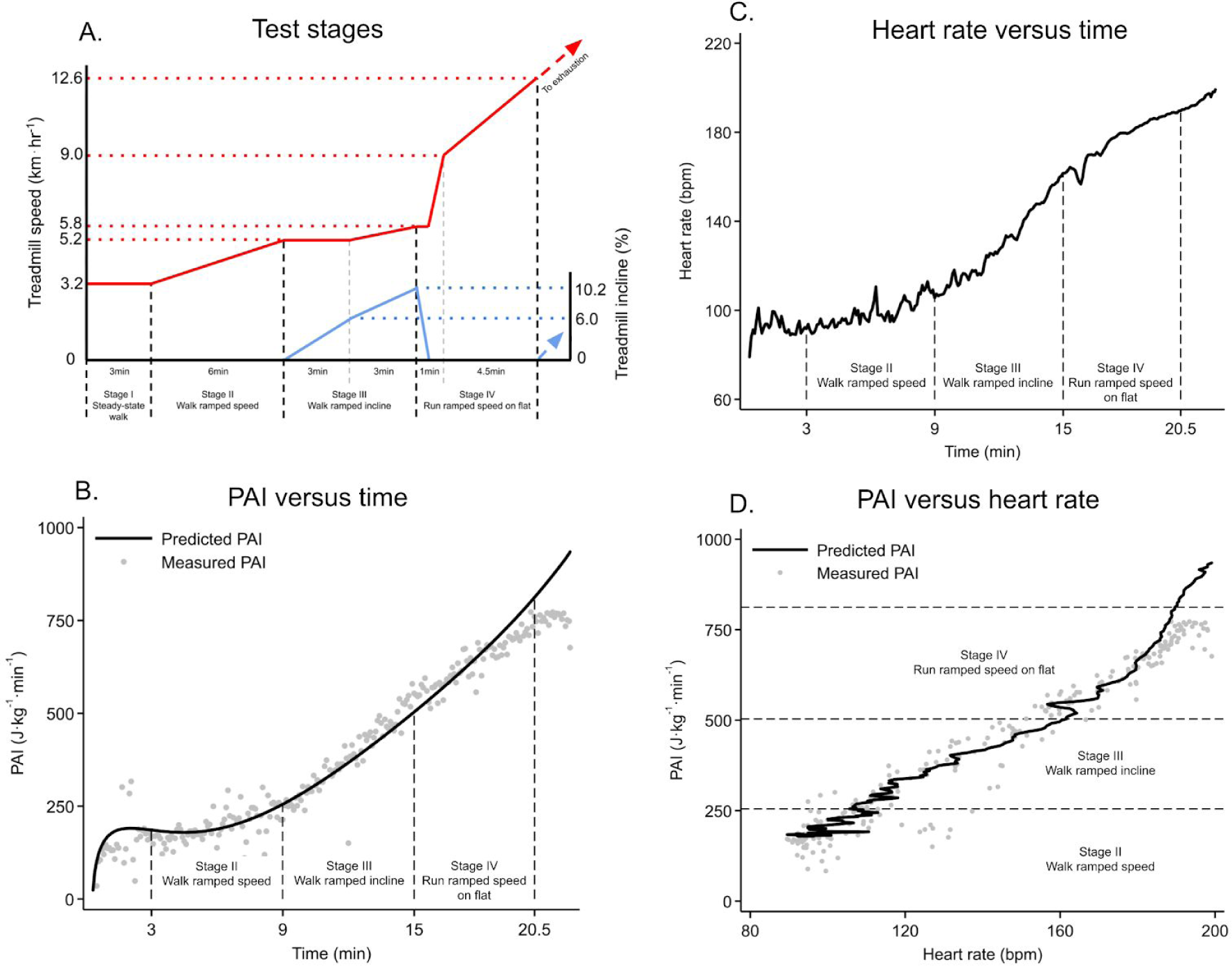
A: Treadmill speed (solid red lines) and incline (solid blue lines) over different stages (time periods between dashed black vertical lines) of the Cambridge Ramped Treadmill Test (CRTT). Dotted red horizontal lines represent treadmill speeds. Dotted blue horizontal lines represent treadmill inclines. Dashed vertical grey lines divide subphases within stages III and IV. To measure VO_2_ max in this study, stage IV was extended (dashed red and blue arrowed lines) until exhaustion was achieved. B: Exemplar data of predicted (solid line) and directly measured (grey dots) physical activity intensity (PAI) across stages II-IV. C: Exemplar data of heart rate across stages II-IV. D: Exemplar data demonstrating PAI-to-heart rate relationship across stages II-IV.

### Measurement of VO_2_ max

The CRTT was extended with an additional stage to allow direct measurement of VO_2_ max in this study. After Stage IV of the original protocol, treadmill speed was increased by 0.25 km·h^-1^ and incline by 0.5% every 15s until exhaustion was reached. Breath-by-breath values of VO_2_ were averaged in 15s epochs, filtering out the highest and lowest breath values, and VO_2_ max was computed as the average of the two largest VO_2_ values in the last 45s of the test. The test was terminated if one of the following three criteria were satisfied: 1) the participant wanted to stop despite verbal encouragement; 2) participant indication of angina, light-headedness, or nausea; and 3) failure of the testing equipment. In analysis, VO_2_ max was considered reached if two of the four following criteria were achieved: 1) respiratory exchange ratio value > 1.2; 2) leveling-off in VO_2_ (< 2.5 ml O_2_·kg^-1^·min^-1^ change) despite an increase in WR; 3) leveling-off in HR (< 3bpm per min) despite an increase in WR; and 4) reaching 100% of the participant’s age-predicted HRmax ^5^.

### VO_2_ max prediction methods

We examined the validity of predicted VO_2_ max values computed from three prediction methods: 1) a HR-to-WR linear regression method using data from CRTT stages II, III, and IV; 2) a single-point (low-point) walk-test calibration method using data from CRTT stage I (minutes 2:30 to 3:00); and 3) a single-point (high-point) walk-test calibration method using data from CRTT stage II (minutes 8:00 to 10:00). The two latter models were derived in different samples of individuals ^12,13^. All methods effectively model the linear relationship between HR and net oxygen cost; this was extrapolated to HRmax and an estimate of resting energy expenditure was added to predict VO_2_ max. Multiple VO_2_ max predictions were computed per participant to examine the validity of different combinations of test endpoints, extrapolation endpoints, and resting energy expenditure estimates. The processes used to predict VO_2_ max values for each method are described in Supplementary Figures 1, 2, and 3.

**Figure 2.**
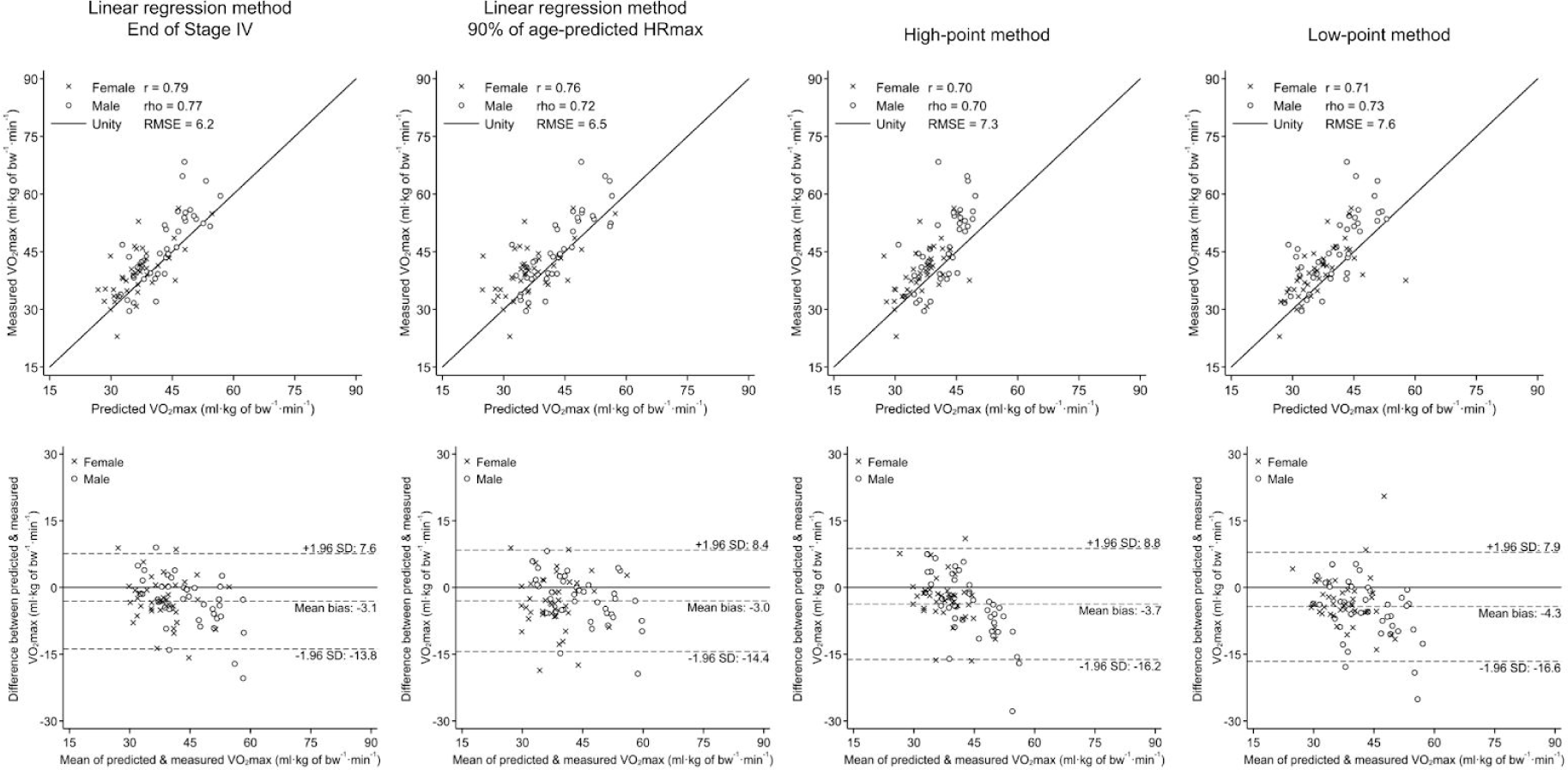
Scatterplots (top row) and Bland-Altman plots (bottom row) demonstrating agreement between predicted and directly measured VO_2_ max per kilogram whole-body mass (bw). Columns show agreement results for selected field-use scenarios. Different test endpoints are demonstrated for the linear regression method. Age-predicted HRmax was used as the extrapolation endpoint for all plots shown. r: Pearson’s correlation coefficient, rho: Spearman’s rank correlation coefficient, RMSE: Root mean squared error

**Figure 3.**
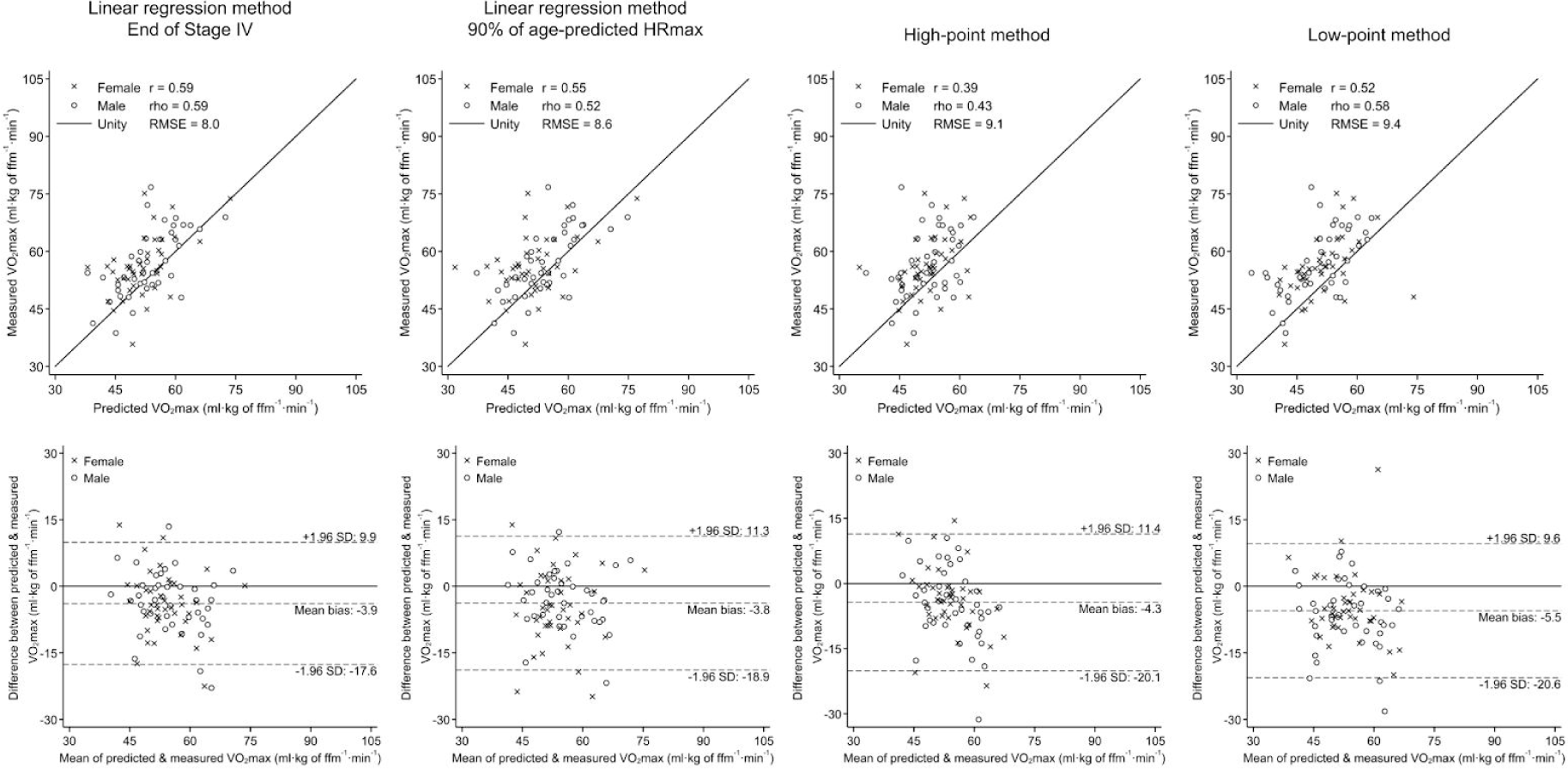
Scatterplots (top row) and Bland-Altman plots (bottom row) demonstrating agreement between predicted and directly measured VO_2_ max per kilogram fat-free mass (ffm). Columns show agreement results for selected field-use scenarios. Different test endpoints are demonstrated for the linear regression method. Age-predicted HRmax was used as the extrapolation endpoint for all plots shown. r: Pearson’s correlation coefficient, rho: Spearman’s rank correlation coefficient, RMSE: Root mean squared error

## Statistical analyses

We report the mean age-predicted HRmax percentage achieved at each CRTT stage. Correlations between predicted and directly measured VO_2_ max were quantified using Pearson’s *r* and Spearman’s *rho*. Bias was computed as the difference between predicted and directly measured VO_2_ max. One-sample t-tests were performed to determine whether mean biases were statistically significantly different from zero. Differential bias was examined by sex and across combinations of test endpoints (CRTT stages and percentages of age-predicted HRmax). Sources of prediction error were examined by extrapolation endpoints (age-predicted and measured HRmax) and resting energy expenditure (REE) estimates (1 MET, Henry (2005)^9^, directly measured ^16^). Prediction precision was expressed as the root mean square error (RMSE). Measurement agreement was visualised using scatterplots and Bland-Altman plots with limits of agreement equal to ± 1.96 standard deviations around the mean bias. Statistical significance was set to 0.05. Statistical analyses were performed with STATA (Version 15.1; StataCorp, College Station, TX).

## Results

Participant characteristics are reported in Table 1. We initially recruited 97 participants (51 females, 46 males). Nine participants were excluded from the present analyses for terminating the treadmill test early and 5 participants for equipment failure, resulting in a final sample of 42 females and 41 males with valid measures of VO_2_ max.

**Table 1.** Participant characteristics.

| Characteristic | Female (n = 42) | Male (n = 41) |
| --- | --- | --- |
| Age (y) | 39.6 ± 12.3 | 41.7 ± 13.5 |
| Height (cm) | 165.1 ± 6.4 | 177.5 ± 6.9 |
| Weight (kg) | 63.4 ± 7.5 | 78.9 ± 11.7 |
| BMI (kg·m<sup>-2</sup>) | 23.2 ± 2.6 | 25.1 ± 3.6 |
| Fat free mass (kg) | 44.5 ± 3.4 | 62.9 ± 6.4 |
| RHR (bpm) | 60.9 ± 8.5 | 60.1 ± 10.7 |
| Measured HRmax (bpm) | 186.3 ± 11.4 | 186.7 ± 10.6 |
| Age-predicted HRmax (bpm) | 180.3 ± 8.6 | 178.8 ± 9.4 |
| Measured REE (J·kg<sup>-1</sup>·min<sup>-1</sup>) | 59.4 ± 7.7 | 60.6 ± 8.4 |
| Predicted REE (J·kg<sup>-1</sup>·min<sup>-1</sup>) | 62.6 ± 4.4 | 64.4 ± 4.4 |
| VO<sub>2</sub>max per kg bodyweight (ml O<sub>2</sub>·kg<sup>-1</sup>·min<sup>-1</sup>) | 39.7 ± 6.7 | 46.0 ± 9.5 |
| VO<sub>2</sub>max per kg fat-free mass (ml O<sub>2</sub>·kg<sup>-1</sup>·min<sup>-1</sup>) | 56.0 ± 7.9 | 56.8 ± 8.7 |

BMI: body mass index; RHR: resting heart rate; HRmax: maximal heart rate; REE: Resting energy expenditure; VO<sub>2</sub>max: maximal oxygen consumption; Age-predicted HRmax = 208 - 0.7 · age(y); Predicted REE was computed according to Henry (2005)

The relative intensity of CRTT stages I, II, III and IV were 34 ± 8, 41 ± 9, 72 ± 15, and 100 ± 12% age-predicted HRmax, respectively. Table 2 demonstrates levels of agreement between predicted and directly measured VO_2_ max when scaled by whole-body mass, using different prediction methods, and across different test endpoints. For this table, the extrapolation endpoint was age-predicted HRmax and resting energy expenditure was computed according to the equations by Henry (2005) ^9^, reflecting the approach used in most field-use scenarios. The linear regression method demonstrated good levels of agreement with measured VO_2_ max when the end of stages III and IV were used as test endpoints (Pearson’s *r* range: 0.67 to 0.79; mean bias range: −3.1 to −2.5 ml O_2_·kg^-1^·min^-1^) or when the attainment of 80% to 100% of the participants age-predicted HRmax was used as a test endpoint (Pearson’s *r* range: 0.72 to 0.79; mean bias range: −3.0 to −2.6 ml O_2_·kg^-1^·min^-1^). All other test endpoint criteria examined resulted in worse levels of agreement for the regression-based methods. Predicted VO_2_ max values from the low-point method were correlated to directly measured VO_2_ max (Pearson’s *r*: 0.71) but with statistically significant underestimation bias (−4.3 ± 6.2 ml O_2_·kg^-1^·min^-1^; *p* < 0.01). Similarly, predicted VO_2_ max values from the high-point method were also correlated to directly measured VO_2_ max (Pearson’s *r*: 0.69) and with a smaller but still statistically significant underestimation bias (−3.5 ± 6.4 ml O_2_·kg^-1^·min^-1^; *p* < 0.01). Scatterplots and Bland-Altman plots in Figure 2 demonstrate levels of agreement for test endpoints in four selected field-use scenarios. Analogous agreement results but with VO_2_ max scaled by fat-free mass instead of whole-body mass are shown in Table 2 and Figure 3. In comparison to whole-body mass scaling, scaling by fat-free mass resulted in agreement results with less precision but similar bias values.

**Table 2.** Agreement between predicted and directly measured VO_2_ max per kilogram whole-body mass across prediction methods. Values were computed using the participant’s age-predicted HRmax as the extrapolation endpoint and resting energy expenditure values according to the equations by Henry (2005) ^9^. Different test endpoints were used for the linear regression method to simulate field-use scenarios.

| VO<sub>2</sub>max prediction method | Test endpoint | RMSE | Pearson's *r* | Spearman's *ρ* | Pooled bias | Female bias | Male bias |
| --- | --- | --- | --- | --- | --- | --- | --- |
| Linear regression method | Stage IV | 6.2 | 0.79 | 0.77 | -3.1 ± 5.5* | -2.9 ± 5.1* | -3.3 ± 5.9* |
|  | Stage III | 7.3 | 0.67 | 0.64 | -2.5 ± 6.9* | -3.6 ± 6.5* | -1.3 ± 7.2 |
|  | Stage II | 16.9 | 0.11 | 0.17 | -14.2 ± 9.2* | -13.4 ± 7.6* | -15.2 ± 10.8* |
|  | 100% age-predicted HRmax | 6.0 | 0.79 | 0.76 | -2.6 ± 5.5* | -2.9 ± 5.7* | -2.2 ± 5.3* |
|  | 90% age-predicted HRmax | 6.5 | 0.76 | 0.72 | -3.0 ± 5.8* | -3.4 ± 6.1* | -2.6 ± 5.5* |
|  | 80% age-predicted HRmax | 6.8 | 0.72 | 0.67 | -2.9 ± 6.2* | -3.5 ± 6.3* | -2.3 ± 6.1* |
|  | 70% age-predicted HRmax | 7.4 | 0.64 | 0.63 | -2.3 ± 7.0* | -3.2 ± 6.8* | -1.5 ± 7.3 |
|  | 60% age-predicted HRmax | 8.1 | 0.58 | 0.58 | -1.6 ± 8.0 | -3.0 ± 8.0* | -0.1 ± 7.9 |
|  | 50% age-predicted HRmax | 10.0 | 0.57 | 0.55 | -3.0 ± 9.6* | -6.0 ± 9.4* | 0.1 ± 8.9 |
| High-point method | Stage II | 7.3 | 0.69 | 0.69 | -3.5 ± 6.4* | -2.8 ± 5.4* | -4.3 ± 7.3* |
| Low-point method | Stage I | 7.6 | 0.71 | 0.73 | -4.3 ± 6.2* | -3.2 ± 5.7* | -5.5 ± 6.6* |

Age-predicted HRmax = 208 - 0.7 · age(y); RMSE: root mean squared error; Bias values are mean ± standard deviation ml O<sub>2</sub>·kg<sup>-1</sup>·min<sup>-1</sup>; \*: One-sample t-test *p* < 0.05

We examined agreement between different combinations of prediction methods, test endpoints, extrapolation endpoints, and resting energy expenditure estimates when scaled by whole-body mass (Supplementary Tables 1-4) and fat-free mass (Supplementary Tables 5-8), allowing quantification of the relative contribution of uncertainty from the various components in each model. Compared with measured HRmax (Supplementary Tables 1 and 5), the largest source of prediction bias across methods stem from the use of age-predicted HRmax as the extrapolation endpoint (Supplementary Tables 2 and 6). Differences in accuracy and precision across REE estimation methods were negligible (Supplementary Tables 3 and 7). For the linear regression method (Supplementary Tables 4 and 8), prediction precision declined across test endpoints (50% through 100% age-predicted HRmax; Stage II through IV) while bias values remained relatively stable except for when Stage II was used as an endpoint, resulting in much larger bias.

**Table 3.** Agreement between predicted and directly measured VO_2_ max per kilogram fat-free mass across prediction methods. Values were computed using the participant’s age-predicted HRmax as the extrapolation endpoint and resting energy expenditure values according to the equations by Henry (2005) ^9^. Different test endpoints were used for the linear regression method to simulate field-use scenarios.

| $\text{VO}_2\text{max}$ prediction method | Test endpoint | RMSE | Pearson's $r$ | Spearman's $\rho$ | Pooled bias | Female bias | Male bias |
| --- | --- | --- | --- | --- | --- | --- | --- |
| Linear regression method | Stage IV | 8.0 | 0.59 | 0.59 | $-3.9 \pm 7.0^*$ | $-4.1 \pm 7.1^*$ | $-3.7 \pm 7.0^*$ |
| | Stage III | 9.6 | 0.38 | 0.40 | $-3.2 \pm 9.1^*$ | $-5.1 \pm 9.1^*$ | $-1.2 \pm 8.8$ |
| | Stage II | 22.2 | -0.15 | -0.13 | $-18.9 \pm 11.7^*$ | $-18.8 \pm 10.2^*$ | $-19.1 \pm 13.2^*$ |
| | 100% age-predicted HRmax | 7.9 | 0.60 | 0.59 | $-3.3 \pm 7.3^*$ | $-4.2 \pm 8.0^*$ | $-2.4 \pm 6.5^*$ |
| | 90% age-predicted HRmax | 8.6 | 0.55 | 0.52 | $-3.8 \pm 7.7^*$ | $-4.8 \pm 8.6^*$ | $-2.8 \pm 6.7^*$ |
| | 80% age-predicted HRmax | 9.0 | 0.46 | 0.43 | $-3.7 \pm 8.2^*$ | $-4.9 \pm 8.9^*$ | $-2.5 \pm 7.4^*$ |
| | 70% age-predicted HRmax | 9.7 | 0.33 | 0.34 | $-2.9 \pm 9.3^*$ | $-4.4 \pm 9.5^*$ | $-1.4 \pm 8.9$ |
| | 60% age-predicted HRmax | 10.8 | 0.25 | 0.29 | $-2.0 \pm 10.7$ | $-4.3 \pm 11.2^*$ | $0.4 \pm 9.8$ |
| | 50% age-predicted HRmax | 13.7 | 0.30 | 0.28 | $-4.1 \pm 13.1^*$ | $-8.7 \pm 13.6^*$ | $0.6 \pm 11.0$ |
| High-point method | Stage II | 9.1 | 0.39 | 0.43 | $-4.3 \pm 8.0^*$ | $-3.8 \pm 7.4^*$ | $-4.8 \pm 8.7^*$ |
| Low-point method | Stage I | 9.4 | 0.52 | 0.58 | $-5.5 \pm 7.7^*$ | $-4.5 \pm 7.7^*$ | $-6.5 \pm 7.7^*$ |

Age-predicted HRmax = $208 - 0.7 \cdot \text{age}(y)$ ; RMSE: root mean squared error; Bias values are mean ± standard deviation $\text{ml O}_2 \cdot \text{kg}^{-1} \cdot \text{min}^{-1}$ ; \*: One-sample t-test $p < 0.05$

## Discussion

We examined the validity of two overall approaches to VO_2_ max prediction from HR response to the CRTT: 1) a HR-to-WR linear regression method using individual-level HR response data across multiple CRTT stages; and 2) single-point walk-test calibration methods using the level of HR reached at the end of either CRTT stage I (low-point method) or stage II (high-point method). For the linear regression method, prediction bias ranged from −3.0 to −1.6 ml O_2_·kg^-1^·min^-1^ and Pearson’s *r* ranged from 0.57 to 0.79 for endpoints at different percentages of age-predicted HRmax. Prediction validity was similar when stages III and IV were used as endpoints, but poorer when stage II was used. Interestingly, the low- and high-point methods elicited results comparable to those obtained with the linear regression method. Our findings demonstrate that VO_2_ max can be predicted from the CRTT with acceptable validity across several epidemiological scenarios.

We examined the validity of VO_2_ max predictions at a variety of CRTT endpoint criteria (i.e. percentages of age-predicted HRmax; end of CRTT stages) which reflect field-use scenarios commonly encountered in population-based studies. Terminating testing at a percentage of age-predicted HRmax offers researchers flexibility to tailor the test to the participant’s estimated fitness level whilst keeping risk of adverse events manageable; this increases the proportion of participants who can safely be tested in a given cohort. Nevertheless, age-predicted HRmax has large uncertainty; the standard deviation of the difference between age-predicted and measured HRmax is about 11bpm. Thus, at a given percentage of age-predicted HRmax, participants with lower fitness may exercise at or near maximum capacity while those with higher fitness are yet to be exercising over an adequate range of HR response to allow estimation of the HR-to-WR relationship ^2^. Age-predicted HRmax can disagree considerably with measured HRmax in certain clinical populations, for example those taking cardioactive medications like beta and calcium-channel blockers. Terminating testing upon stage completion would standardise the exercise load across participants, but may be too restrictive for most population-based research. Whatever standard one adopts, we offer the following recommendations and procedures based on our validity findings: 1) researchers using age-predicted HRmax as a stopping criterion should set the threshold value between 80% to 90% and derive VO_2_ max predictions using the linear regression approach; the low- or high-point methods should be considered when participants do not achieve at least 80%. 2) researchers electing to terminate testing upon stage completion could use stage IV to achieve the highest levels of agreement; however, this is likely to be near or at maximal exercise capacity for some participants. Therefore, in most field-use scenarios, stage III should be used to maximize participant safety. The low- and high-point prediction methods can be used when terminating testing at stages I or II, respectively. A mixture of methods could be a viable alternative - for example by combining VO_2_ max predictions from both the linear regression and single-stage methods using inverse-variance weighting. Whilst evaluation of the validity of these types of approaches are beyond the scope of this work, our present results do allow estimating the mean bias of simple weighted averages of multiple methods.

The CRTT has several advantages that make it a viable option for population-based research. First of all, the initial 15 min is the common activity of walking which is familiar to most individuals. As a ramped protocol with modest WR increments ^17^, physiologic responses during the CRTT will generally be uniform across a range of WR intensities (See Figure 1 for reference). Therefore, observed health associations with predicted VO_2_ max should be robust to population-based variation in exercise tolerance. For comparison, the Bruce Treadmill Protocol increases WR in large stepwise increments (2-3 METs) every 3 minutes ^18^. This test design may cause VO_2_ max to be overestimated when predicted at low WR increments, which often limits its use to participants with moderate to high exercise tolerance ^19^. Nonetheless, validity of VO_2_ max estimates from Bruce Treadmill test data are comparable to the ones we report here ^20^. Many individualised and standardised ramped treadmill protocols have been designed as alternatives ^21,22^. These alternative approaches often estimate VO_2_ max with multiple regression prediction models that include non-exercise test factors such as age, whole-body mass, and perceived functional ability as independent predictor variables. The absolute validity of multiple regression prediction models for submaximal treadmill tests are mixed, but in general are well correlated to directly measured VO_2_ max ^21,23,24^. When the proportion of explained variance in VO_2_ max is partially or predominantly attributed to non-exercise factors, true longitudinal change in VO_2_ max - independent of change in other factors - tends to be underestimated by the prediction model. Similarly, for the CRTT, longitudinal change in cardiorespiratory fitness should ideally be assessed using the linear regression method as opposed to the low- and high-point methods since these single-stage approaches are more reflective of group-rather than individual-level differences in fitness.

This study has several limitations. Our study consisted of predominantly white healthy UK adults; therefore, additional work is required to assess the validity of VO_2_ max estimates from HR response to the CRTT in children, adolescents, older adults, patient populations, and people of other ethnicities. The prediction methods we examined here do not fully utilise all information collected during a typical deployment of the CRTT, which could provide additional opportunities for VO_2_ max modeling approaches. For example, HR recovery dynamics can be used to estimate VO_2_ max through nonlinear back-extrapolation methods. This could be explored in future research to determine how these dynamics vary with test completion time and whether they may be leveraged in stronger VO_2_ max prediction models. We were unable to utilise that approach here as it would have required each of the methods to be represented by separate physical tests with a recovery phase, rather not just simulated different test durations as we have examined here.

## Conclusions

We demonstrate here that HR response to the Cambridge Ramped Treadmill Test (CRTT) allows predictions of maximal oxygen consumption (VO_2_ max) with acceptable validity across several common field-use scenarios, affirming its utility in population-based research. Future work should determine whether these findings hold in participants with lower fitness levels and morbidity, and to explore the incorporation of recovery HR dynamics in novel modeling approaches that could improve the validity of VO_2_ max estimates.

## Data Availability

The data analysed during the current study are not publicly available because we have not obtained consent for public data sharing from the study participants; however data can be made available for analysis upon reasonable request to the corresponding author.

## Acknowledgements

We are very grateful to the participants who took part in this study. We thank the principal investigators of the Fenland study for allowing us to recruit from this study population, and the functional teams of the MRC Epidemiology Unit (Study Coordination, Field Epidemiology, Anthropometry, Data Management, IT) for supporting the study. We would like to specifically acknowledge Rebekah Steele, Annie Schiff, and Jill Landsbaugh for study coordination and recruitment.

This research was supported by the Medical Research Council Epidemiology Unit (MC_UU_12015/3) and the NIHR Cambridge Biomedical Research Centre (IS-BRC-1215-20014).

